# Performance & Quality Evaluation of Marketed COVID-19 RNA Detection Kits

**DOI:** 10.1101/2020.04.25.20080002

**Authors:** David Surace Kapitula, Zhuopu Jiang, Jianhao Jiang, Jing Zhu, Xiangyong Chen, Claudia Qiao Lin

## Abstract

Compared to other coronaviruses, COVID-19 has a longer incubation period and features asymptomatic infection at a high rate (>25%)^1,2^. Therefore, early detection of infection is the key to early isolation and treatment. Direct detection of the virus itself has advantages over indirect detection. Currently, the most sensitive and commercially validated method for COVID-19 testing is RT-qPCR, designed to detect amplified virus-specific RNA. Reliable testing has proven to be a bottleneck in early diagnosis of virus infection in all countries dealing with the pandemic. Significant performance and quality issues with available testing kits have caused confusion and serious health risks. In order to provide better understanding of the Quality and performance of COVID-19 RNA detection kits on the market, we designed a system to evaluate the specificity (quantitation), sensitivity (LOD) and robustness of the kits using positive RNA and pseudovirus controls based on COVID-19 genomic sequence^3,4^. We evaluated 8 Nucleic Acid qPCR Kits approved in China, some of which are also approved in the US and EU. Our study showed that half of these 8 kits lack 1:1 linear relationship for virus RNA copy: qPCR signal. Of the 4 with linear response, 2 demonstrated sensitivity at 1 Copy viral RNA/Reaction, suitable for early detection of virus infection. Furthermore, we established the best RNA extraction, handling and qPCR procedures allowing highly sensitive and consistent performance using BGI qPCR kits. Our study provides an effective method to assess and compare performance quality of all COVID-19 nucleic acid testing kits, *globally*.

**Significance Statement:** Testing for COVID-19 has been a critical topic in the pandemic management since the first outbreak reported in China, and now globally. Despite of focused efforts from global biomedical industries and regulatory authorities, testing tools currently available on the market are not satisfying the huge and most important needs for virus control, which is specific, sensitive, affordable, and commercially viable early diagnosis of infected populations. We have designed an experimental system to assess and compare all nucleic acid-based COVID-19 testing kits from quality control perspectives. The results reported here demonstrate the suitability of using our system as an objective QC system for all commercial kits, including any future kits. We also identified the best testing method using commercially available reagents.

## Introduction

Since the novel Coronavirus (COVID-19) was first identified in Wuhan, China, December, 2019, Chinese scientific communities have been making important contributions to the understanding of this virus from genomic sequence to protein structures, infection mechanism to clinical strategies and post-mortem pathology, etc.^4–6^ Most significantly, Chinese epidemiologists have done an impressive job in contact tracing, testing, quarantining of all potentially infected people in China, the majority of whom came from Wuhan, and implementing a strict lock-down, effectively minimizing further spread to the rest of China.

One of the biggest challenges in effective control of the COVID-19 outbreak is the fact that compared to similar viruses, it is more contagious due to its environmental stability; having longer incubation, higher rate of non-symptomatic infections as demonstrated from multiple independent sources^7^, making early detection of virus infection one of the most critical tactics in the battle against the virus. Recognizing the criticality and need early, the China Center for Drug Evaluation (CDE) announced a “Green Channel” for reviewing and approving COVID-19 testing kits and other bio-medical interventions such as clinical trial drugs in January 2020. At the time of submission, 23 diagnostic kits have been approved in China, of which 8 are based on quantitative Polymerase Chain Reaction (qPCR) using COVID-19 viral RNA sequence as templates and fluorescence detection. *See Table 4 in the Tables section*.

These detection kits provided the Chinese CDC and related clinics the ability to test for the presence of infection. However, significant technical and performance quality issues with these testing kits have become evident. In China, the reliability of detecting COVID-19 using approved qPCR methods have been reported by first line healthcare professionals to range from 35–50%^8^. This situation resulted in the initial under-reporting of COVID-19 infected cases in China, which in turn triggered the change of diagnostic criteria by China CDC on Feb 11 from solely the RNA test result to relying on a combination of clinical symptoms including lung CT scan and RNA test^9^. These issues are now becoming evident in the pandemic management in the world. Continued global spread of the epidemic highlights the continual and present demand for sensitive and reliable virus detection without exception, including countries considered as leading bio-medical countries (i.e. EU nations and the US). Notably, the initial failure in quality virus testing kits distributed by US CDC is viewed as a critical mistake in the management of the crises by the US government resulting in compromised ability to test and trace the earlier infected population clusters before they spread to wider populations through community contacts. The failure in providing a sufficient amount of reliable quality testing tools has since been identified as a critical factor in the rapid spread of the virus in many parts of the world which is paralyzing healthcare systems and claiming hundreds of thousands of lives so far.

Our study aims at providing objective evaluation and comparison of the quality and performance characteristics of 8 of the currently marketed COVID-19 nucleic acid detection kits in China based on qPCR and fluorescence detection. The parameters assessed include specificity (quantitation) of the testing kit to COVID-19 gene sequences, sensitivity (LOD), as well as the RNA extraction reagents and processing methodologies.

## Results

### Stage 1 (*see Materials and Methods*): Quantitation Assessment and Amplicon Mapping

The data (CT#) in Table 1 for each kit was assessed to determine the number of ACs providing a positive result (when performed per manufacturer’s instruction). Kits were reviewed for Positive results for all relevant target genes, as well as non-target. Multiple and Single Positive results for the ORF1a/b regions were used to determine suitability as a robust quantitative analytical method. *See Table 1 in the Tables section*.

**Table 1.** CT Number of 8 Kits Using GP and CSC RNA Controls

| KIT ID | 1 Bio-Germ |  | 2 XABT |  | 3 Daan Gene |  | 4 Sansure |  | 5 LiveRiver |  |  | 6 Maccura |  |  | 7 GeneoDx |  |  | 8 BGI |
| --- | --- | --- | --- | --- | --- | --- | --- | --- | --- | --- | --- | --- | --- | --- | --- | --- | --- | --- |
|  | N-VIC | O-FAM | N-VIC | O-FAM | N-FAM | O-VIC | N-ROX | O-FAM | N-VIC | O-FAM | E-ROX | N-Cy5 | O-FAM | E-ROX | N-FAM | O-ROX | E-JOE | O-FAM |
| GP1 | 29.66 | / | 30.17 | / | 35.06 | / | 33.32 | / | 30.81 | 37 | 31.4 | 31.9 | 30.9 | 31.62 | / | / | / | / |
| GP2 | 29.91 | / | 30.43 | / | 32.72 | 36.39 | 32.32 | / | 30.45 | / | 30.7 | 30.7 | / | 30.16 | 32.8 | 37 | 32.6 | 28.91 |
| GP3 | 29.89 | / | 30.77 | / | 33.31 | 34.82 | 32.76 | / | 30.61 | 29.28 | 30.9 | 31.4 | / | 31.78 | / | / | 38.9 | / |
| GP4 | 29.27 | / | 30.84 | / | 32.48 | 34.02 | 31.25 | 30.98 | 29.85 | 20.8 | 30.9 | 30.5 | / | 31.19 | 35 | / | 36.4 | / |
| GP5 | 7.73 | 7.88 | 8.1 | 7.45 | 7.03 | 27.92 | 7.52 | / | 6.76 | 34.95 | 6.77 | 6.85 | 24.5 | 6.31 | 7.83 | 10 | 8.82 | / |
| GP6 | 28.06 | / | 30.73 | / | 32.19 | 32.28 | 31.96 | / | 30.65 | / | 30.8 | 31.5 | 25.7 | 30.22 | 33.2 | 39 | 33.3 | / |
| GP7 | 29.11 | / | 30.79 | / | 33.57 | 38.95 | 34.68 | / | 31.79 | 37.9 | 32.5 | 31.4 | / | 31.41 | 34.8 | / | 34.8 | / |
| GP8 | 28.47 | / | 30.82 | / | 31.3 | 27.33 | 32.56 | / | 29.57 | / | 30.1 | 30.8 | / | 30.38 | 34.2 | 38 | 34.2 | / |
| CSC | 31.14 | / | 31.44 | / | 24.12 | 16.46 | / | / | 32.9 | / | 32.9 | 29.5 | 26.9 | 32.71 | 32.7 | / | 34.5 | / |
| Pos. | 22.76 | 23.9 | 19.55 | 20.02 | 32.96 | 30.75 | * | * | 27.47 | 25.57 | 27.9 | 32.6 | 37.6 | / | 28.4 | 30 | 29 | 26.88 |
| Neg. | / | / | / | / | / | / | * | * | / | / | / | / | / | / | / | / | / | / |

\* Sansure Pos. and Neg. control did not test in this assay.
/ represents no amplification

The results show that kits from Shanghai Bio-Germ Medical Technology Co., Ltd, Beijing AB Technology Co., Ltd, Sansure Biotech Co., Ltd and BGI anneal on single area on ORF1a/b. The others amplified from multiple areas of ORF1a/b, indicating non-specific annealing and resulting in a non-linear sensitivity for virus RNA copy number (i.e. not one-to-one). Furthermore, only the kits from Zhongshan Daan Gene Co., Ltd and Maccura Biotech Co., Ltd could amplify from the Chinese National Standard (CSC). *See Table 4 in the Tables section*.

### Stage 2 (*see Materials and Methods*): Reaction Limit of Detection, LOD, (for Quantitative Kits only)

The data (CT#) for each kit was assessed (Table 2) to determine the number of positive results for both H (10 Copies) and L (1 copy) (when performed per manufacturer’s instruction) and/or a single positive result for replicates.

**Table 2.** CT Number of Reactions from Low and High Concentration of Controls

| KIT ID | 1 Bio-Germ |  | 2 XABT |  | 3 Daan Gene |  | 4 Sansure |  | 5 LiveRiver |  |  | 6 Maccura |  |  | 7 GeneoDx |  |  | 8 BGI |
| --- | --- | --- | --- | --- | --- | --- | --- | --- | --- | --- | --- | --- | --- | --- | --- | --- | --- | --- |
|  | N-VIC | O-FAM | N-VIC | O-FAM | N-FAM | O-VIC | N-ROX | O-FAM | N-VIC | O-FAM | E-ROX | N-Cy5 | O-FAM | E-ROX | N-FAM | O-ROX | E-JOE | O-FAM |
| H | / | 36.94 | / | / | / | / | 35.3 | 37.69 | 40.36 | / | 37.88 | 35.7 |  | 34.6 | 39.69 | / | 41.5 | 33.95 |
| L | / | / | / | / | / | / | 37.68 | 38.46 | / | / | / |  |  |  | / | / | / | 35.38 |
| L | / | / | / | / | / | / | 38.53 | 39.7 | / | / | / |  |  |  | / | / | / | 36.77 |
| Pos. | 20 | 19.79 | 21.83 | 21.78 | 29.29 | 27.84 | 28.66 | 33.59 | 26.27 | 23.94 | 24.44 | 32 | 35 | 35.4 | 27.71 | 30.91 | 28.77 | 24.71 |
| Neg. | / | / | / | / | / | / | / | / | / | / | / |  |  |  | / | / | / | / |

/ represents no amplification

Only Kits from Sansure Biotech Co., Ltd and BGI can achieve low concentration (1 copy/reaction), Bio-Germ, LiveRiver and Maccura can achieve 10 Copies/rxn., and the rest were not reactive at or below 10 Copies/rxn. *See Table 2 in the Tables section*.

### Stage 3 (*see Materials and Methods*): Extraction Efficiency and Assay Repeatability

The results (Table 3) show that for the Tiangen and Qiagen extraction kits, RNA extraction and stability are consistent as claimed. The BGI qPCR COVID-19 Detection kit demonstrated highest fidelity in detection of both high and low copies/rxn. *See Table 3 in the Tables section*.

**Table 3.** CT Number of Extractions from Pseudoviruses Followed by RT-qPCR

|  | Sansure |  |  |  |  |  |  |  |  |  |  |  | BGI |  |  |  |  |  |
| --- | --- | --- | --- | --- | --- | --- | --- | --- | --- | --- | --- | --- | --- | --- | --- | --- | --- | --- |
|  | N-ROX |  |  |  |  |  | O-FAM |  |  |  |  |  | O-FAM |  |  |  |  |  |
|  | Tiangen |  | Qiagen |  | Roche |  | Tiangen |  | Qiagen |  | Roche |  | Tiangen |  | Qiagen |  | Roche |  |
|  | Rep-1 | Rep-2 | Rep-1 | Rep-2 | Rep-1 | Rep-2 | Rep-1 | Rep-2 | Rep-1 | Rep-2 | Rep-1 | Rep-2 | Rep-1 | Rep-2 | Rep-1 | Rep-2 | Rep-1 | Rep-2 |
| GP1 | 37.56 | / | / | / | / | / | / | / | 40 | / | / | / | 36.77 | / | 34.96 | / | 35.02 | / |
| GP2 | 37.65 | 37.64 | 40 | 40 | 38.72 | / | / | / | 38.81 | 38.38 | / | / | 36.73 | 37.43 | 37.24 | 37.2 | / | 36.92 |
| GP3 | / | 38.71 | 40 | 40 | / | / | / | 40.3 | 38.24 | 40 | 40.4 | / | 35.66 | / | 35.97 | 37.39 | / | 37.16 |
| GP4 | 36.51 | / | 40 | / | / | / | / | / | 39.35 | 40 | / | / | 36.12 | 37.65 | 36.87 | / | 35.71 | 37.33 |
| GP5 | / | / | / | 40 | 37.99 | / | 40.35 | 40.58 | / | 40 | 40.8 | / | / | 36.8 | / | 36.85 | 35.51 | 36.61 |
| GP6 | / | / | / | 40 | / | / | / | / | 40 | 40 | / | / | / | / | 36.46 | / | / | 35.87 |
| GP7 | / | / | / | 40 | 37.6 | / | / | 40.53 | 40 | / | / | 41.11 | 36.28 | 35.35 | 35.86 | / | 35.23 | / |
| GP8 | 41.72 | 36.72 | 40 | 40 | / | 37.58 | / | 39.97 | 40 | 40 | / | / | 31.22 | 35.76 | 36.72 | / | / | / |
| Extract-Pos. | 27.51 | 27.77 | 30.7 | 30.78 | / | / | 30.3 | 30.5 | 28.72 | 28.66 | / | / | 26.94 | 26.73 | 26.67 | 26.6 | 27.49 | 27.79 |
| Extract-Neg. | / | / | / | / | / | / | / | / | / | / | / | / | / | / | / | / | / | / |

/ represents no amplification

## Discussion

*In Vitro* Diagnostic kits (IVD) are usually approved by regulatory authorities around the world under the category of medical devices. Commercialization of IVDs typically requires 3-5 years of product development, followed by a 1 to 2-year regulatory review period. Evaluation criteria include scientific rationale, technical considerations, product quality and safety, and consistency of performance for the intended purpose and markets including manufacturing capability (supply). The COVID-19 RNA kits approved under EUA had much shorter time for development and regulatory review, in some cases resulting in higher variability and quality issues. These issues often resulted in delays in diagnosis requiring repetitive testing using different testing kits and methods in order to confirm a positive infection as a result of the high rate of “False Negatives” observed in many of the approved testing kits. IVD industry leaders (*i.e*. Roche Diagnostics) are equally affected, although having one of the first COVID-19 IVD approved by the US FDA under EUA, the determined sensitivity, when compared head-to-head with other kits in our study, did not achieve single copy/reaction detectability. ROCHE reagents used in Cobas-SARS-CoV-2^10^ achieved quantitative detection of the viral RNA, consistent with product claim (LOD of 10 copies/reaction) and confirmed in our study (data not shown), while both BGI and Sansure kits demonstrated LODs at 1 copy/reaction. Therefore, the existing RNA testing tools world-wide are at best insufficient in providing consistent performance suitable for reliable early detection and proper isolation, a key-criteria central to the control of the pandemic before an effective vaccine or treatment is available. In addition, lack of reliable correlation of clinical treatment with viral load could also affect decisions on treatment and post-recovery patient management.

Our study provides an elegant design to define the most important performance characteristics of the RNA detection kits for COVID-19, which are specificity (quantitative), sensitivity (LOD), and robustness. Using this approach, all COVID-19 RNA detection kits can be compared with the currently approved kits to aid the decision in regulatory approval as well as clinical applications. Use of the E, N genes and ORF1a/b region of the virus genome as RNA based assay and internal controls in our study takes into consideration that these represent the most likely regions (coding for structural and non-structural proteins, including those essential to virus functions) for Primer and Probe design11. This study design is viable for comparing and screening all new COVID-19 virus detecting reagents in the world to provide objective evaluation of the performance and quality of the new IVD tools.

In conclusion, Jade Biomedical (Suzhou) Co, Ltd has identified the Best Total Solution (BTS) for COVID-19 RNA testing, which involves using Tiangen or Qiagen RNA extraction and BGI qPCR kits. This combination allows reliable detection of viral RNA at 1-10 Copies/reaction, which equates to 95-1000 Copies of viral RNA in 1 mL of test article. Coincidentally, BGI Real-Time Fluorescent RT-PCR kit from China is the only one of its kind to have obtained US FDA and EMA emergency approval so far^12,13^. The Quality Control strategy and reliable performance by Jade Biomedical Laboratories demonstrated in this study will continue to allow contribution to the global need for more reliable and easily accessible COVID-19 testing. Direct clinical validation of our BTS is currently being planned in order to help with the on-going global clinical treatment strategy and intervention development by providing such specific, sensitive, and reliable viral load testing.

## Materials and Methods

### Description of Test Principle and Test Kits in the Study

The COVID-19 RT-PCR test kits are real-time reverse transcriptase polymerase chain reaction (rRT-PCR) tests for the quantitative detection of nucleic acid from SAR-CoV-2. The tests are shown to support detection in upper and lower respiratory specimens (e.g. – nasopharyngeal or oropharyngeal swabs, sputum, lower respiratory tract aspirates, etc.). They are designed with specific primer and probe sets to detect from one to three regions in the COVID-19 genome, 1) Nucleocapsid (N) gene, seq. 28274-29533; 2) ORF1a and ORF1b – various functional proteins, seq. 266-21555; and 3) Envelope Protein (E) gene, seq. 26245-26472 (Table 4).

**Table 4.** Real-Time qPCR Primer and Probe Targets# for Test Reagents^15–22^

| Kit Identifier Number | Manufacturer | Registration No. (China) | Target Gene(s)# |
| --- | --- | --- | --- |
| 1 | Shanghai Bio-Germ Medical Technology Co., Ltd. | 20203400065 | N, ORF1a/b |
| 2 | Beijing AB Technology Co., Ltd. | 20203400179 |  |
| 3 | Zhongshan Daangene Co., Ltd. | 20203400063 |  |
| 4 | Sansure Biotech Co., Ltd. | 20203400064 |  |
| 5 | Shanghai ZJ Bio-Tech Co., Ltd. | 20203400057 | N, ORF1a/b, E |
| 6 | Maccura Biotech Co., Ltd. | 20203400184 |  |
| 7 | Shanghai Geneodox Co., Ltd. | 20203400058 |  |
| 8 | BGI | 20203440060 | ORF1a/b |

Quantitative real-time qPCR is performed using the Roche LightCycler 480 real-time PCR System (LC480)14. RNA isolated from specimens is reverse transcribed to cDNA and subsequently amplified using one of the kits in Table 4. The specific primer target(s) (amplicon) is/are amplified via Reverse Transcription (RT), denaturation, annealing, and extension of the primers followed by detection of the fluorescent reporter dye(s), per manufacturer’s instructions. *See Table 4 in the Tables section*^15–22^.

To enable the objective performance evaluation of the kits, and due to non-disclosure by some kit manufacturers, a total of 8 synthetic RNA controls (AC) were synthesized, purified, and individually packaged into Lentivirus for RNA extraction studies, Pseudovirus (IC). The sequence design of these controls was based on RNA length supported by the Lentivirus, (Refer to Table 5 below for sequence specifics). Furthermore, to facilitate the ability to support single and multi-plex assay designs, each of the controls also contained the full-length N and E genes, as well as a specific sequence with partial overlap of the ORF1a/b sequence on both the 3’ and 5’ ends (Table 5). The partial overlap was designed to provide sufficient detectability in the event the primers/probe combinations of the commercial kits were designed to anneal near the boundaries of the included ORF1a/b segment. In addition, the Chinese National Standard, a synthetic RNA (CSC) similar in construct of the AC was also included as a screening element. *See Table 5 in the Tables section*.

**Table 5.** Assay Control (AC) and Internal Control (IC) Gene Sequence Information

| Control Sequence Code | E Gene<sup>3</sup> | N Gene<sup>3</sup> | ORF1a/b Region<sup>3</sup> |
| --- | --- | --- | --- |
| GP1 | 26245-26472 | 28274-29533 | 266-3006 |
| GP2 |  |  | 2856-5667 |
| GP3 |  |  | 558-8329 |
| GP4 |  |  | 8180-10990 |
| GP5 |  |  | 10841-13651 |
| GP6 |  |  | 13502-16312 |
| GP7 |  |  | 16163-18973 |
| GP8 |  |  | 18824-21555 |
| CSC | 26245-26472 | 28274-29533 | 14911-15910 |

### Plasmids

SARS-nCoV-2 genomic sequence (MN908947.3)^3^ was acquired from NCBI. All gene fragments used in this study were assembled by overlapping synthesized oligonucleotides. The whole ORF1a/b sequence was divided evenly into 8 fragments with 150bp overlaps in between each other. Genes E and N were added to the 3’ end of each ORF1a/b fragments, then cloned into RNA producing plasmid pUC57 (Thermo Scientific, Carlsbad, CA) or Lentivirus transfer plasmid pLEX-MCS (Addgene, Cambridge, MA).

### *In-Vitro* Transcription (IVT)

To prepare DNA templates for IVT, the synthesized SARS-nCoV-2 fragments, flanked by T7 promoter at 5’ end and 30 poly(A)s at 3’ end, were amplified by Phanta Max Super-Fidelity DNA Polymerase (Vazyme, Nanjing). IVT of RNAs were using RNA TranscriptAid T7 High Yield Transcription Kit (Thermo Scientific, Carlsbad, CA) with extended DNase I treatment (1 hour). Then RNAs were purified by Monarch RNA Cleanup Kit (NEB, Ipswich, MA) and diluted to 1 × 10^6^ copies/mL based on OD260 readout by Nanodrop One (Thermo Scientific, Carlsbad, CA).

### Virus Production and Titration

For Lentivirus packing, SARS-nCoV-2 sequence containing pLEX-MCS (Addgene, Cambridge, MA), psPAX2 (Addgene, Cambridge, MA) and pVSV-G (Addgene, Cambridge, MA) were co-transfected by RNAi-Mate (GenePharma, Suzhou) into HEK293T cells. Virus was harvested from culture medium 72 hours after transfection by filtration through 0.45μm filters (Merck Millipore, Nottingham, UK). The flow through was moved into 100kd Centrifugal Filters (Merck Millipore, Nottingham, UK) and spun @8000rpm in 4°C for virus concentration. Virus was treated by DNase I (Thermo Scientific, Carlsbad, CA) for 30 minutes before RT-qPCR titration.

To quantify genomic RNA in SARS-nCoV-2 fragment replaced Lentivirus, the virus was lysed by Ezol (GenePharma, Suzhou) and extracted with chloroform. Purified RNA was then reverse-transcribed into DNA by M-MLV Reverse Transcriptase (Thermo Scientific, Carlsbad, CA). RT-qPCR reactions using Primer set S-LV5-FO-3 (5’-AGAGCGTCAGTATTAAGCGGG-3’) and S-LV5-RE-3 (5’-TGTTTCTAACAGGCCAGGATT-3’) were performed in Applied Biosystems 7500 Real-Time PCR System (Thermo Scientific, Carlsbad, CA). Copy number of virus was calculated by standard curve using plasmid DNA. Real-time fluorescence quantitative general reagent (Cat# GMRS-001 GenePharma, Shanghai) was used to detect copy number of RNAs.

### Experimental Design

The kits were evaluated using a stepwise three-stage approach. All assays were performed in accord with the manufacturer’s instruction using the LightCycler (Roche) (adjusted for color compensation for the specific Reporter Dye(s).

### Stage 1: Quantitation

Performed to verify the ability of the assay to be used for quantitative analysis and to determine the target region of the primer(s) and probe(s) for each manufacturer’s kit. Only AC (RNA) was used for this stage of the study. Each assay included kit provided Positive and Negative Controls. As all commercial kits used have a claimed LOD of ≤ 1000 copies/reaction), Master Mix was prepared to which *1000 copies of AC were added, (note the AC, GP5 was spiked at 10^8^ copies). Additionally, separate samples using CSC were run in parallel.

### Stage 2: Reaction Limit of Detection

Those kits demonstrated suitable for quantitative analysis (from Stage 1) were assayed with the relevant single ACs using two concentrations of spiked AC. Each assay included kit provided Positive and Negative Controls. Master Mix was prepared to which 2 copies/µl (@10 copies/rxn., (H) and 0.2 copies/µl @1 copy/rxn., (L) of identified (Stage 1) AC were added with samples being run in duplicate.

### Stage 3: Extraction Efficiency and Assay Repeatability

For those kits with an established LOD at 1 Copy/rxn, the robustness of the testing was evaluated using 3 available RNA extraction kits, Tiangen^23^, Qiagen^24^, and Roche ^25^. This stage was performed to identify the best combination of extraction reagent, method, storage and processing conditions, and RT-PCR reagents providing the most sensitive and reliable detection of low levels of COVID-19 infection, termed “Best Total Solution” (BTS) COVID-19 RNA Detection Method.

All extractions were performed in accord with the manufacturer’s instructions. The relevant IC (from Stage 1) was used for each of the kits assessed. Eight (8) samples were individually extracted for each kit. A Positive and Negative Control (included in the kits) were also prepared. Pseudovirus RNA Extraction was performed using IC @1000 Copy/mL Based on 90% extraction efficiency per manufacturer’s claims, final RNA Extraction concentrations were assessed to be ~2.1 copy/µl. 5 µl of extracted solution (10 Copies/rxn.) were used for all assays and all samples were run in duplicate.

## Data Availability

I certify that all data referred to in the manuscript are available.

